# Reimagining COVID Vaccine Distribution: Reflecting on Waste and Equity

**DOI:** 10.1101/2024.10.04.24314854

**Authors:** Scott Greenhalgh, Maria L. Alva, Jim Phuong

## Abstract

**Importance:** Efficient distribution and administration of vaccines are critical to preventing unnecessary morbidity and mortality. We assess the distribution, uptake, and wastage of COVID-19 vaccine doses across the U.S., providing insights for optimizing future vaccination distribution strategies.

**Objective:** We evaluate the distribution, uptake, and wastage of COVID-19 vaccine doses in the U.S. Specifically, we quantify the impact of limiting vaccine wastage and illustrate incidence and deaths averted under two targets set by the Global Alliance for Vaccines and Immunization (GAVI).

**Design and Setting:** We obtained COVID-19 vaccine doses administered by location and wastage data from jurisdictions, pharmacies, and federal entities from the Centers for Disease Control and Prevention through a Freedom of Information Act. From this data, a retrospective analysis covering the period from December 2020 to October 2022 involving 761 million vaccine doses distributed across all counties and states in the U.S. We estimate the proportion of vaccines wasted, and then incidence and deaths averted had adherence to GAVI waste targets occurred to inform on the quality of the national vaccination effort and identify potential regions for improvement.

**Exposure:** Vaccine uptake and waste vary substantially across states, as measured by doses administered per capita. GAVI targets of 25% and 15% vaccine waste serve as benchmarks for assessing the impact of potential improvements in vaccine distribution and acceptance.

**Main outcomes and measures:** The identification of within and across-state variation in COVID-19 vaccine waste relative to GAVI targets and their implications on morbidity and mortality.

**Results:** Among the 761 million distributed doses, only 600 million were administered, resulting in a national average of 1.8 doses per capita. Substantial regional disparities were observed, with the District of Columbia reaching 2.5 doses per capita and Alabama lagging at 1.3 doses per capita. Thirty states exceeded the GAVI 15% vaccine waste target, corresponding to 64.2 million unused doses. Meeting the 15% target would have averted 29,669,318 incidences and 6,468 deaths.

**Conclusion and relevance:** Addressing the causes of county-level variations and targeting states with below-average vaccine hesitancy and above-target vaccine waste would likely maximize future vaccine distribution efforts and minimize wastage-related losses. This strategy highlights an avenue for improving future vaccine distribution policy.

**KEY POINTS:** 

**Question:** In what areas of domestic vaccine allocation could improvements be made to reduce vaccine waste? What impact could reducing vaccine waste have had on lowering both COVID-19 incidence rates and mortality rates?

**Findings:** Between December 2020 and October 2022, the U.S. wasted approximately 25.4 million COVID-19 vaccine doses. Reducing waste to under 25% could have averted 1.3 million COVID-19 cases and an estimated 1,570 deaths over that period. Waste was associated with hesitancy, rurality, and prevalent political affiliation.

**Meaning:** This counterfactual exercise underscores the importance of addressing vaccine wastage to mitigate COVID-19 incidence and its associated fatalities.

## INTRODUCTION

The emergency use of COVID-19 vaccines was authorized in December 2020^1^ and distributed by the Federal government based on the size of each state’s adult population without copays until June 2021.^2^ Pfizer, Moderna, and Janssen distribution in the U.S. rolled out across multiple phases to multiple priority groups to mitigate severe illness and protect individuals whose roles are critical to society’s health, and functionality. As of May 2023, 81.4% of the U.S. population has been vaccinated at least once, with 69.5% completing the primary series of vaccinations.^3^ Despite this national vaccination level, vaccination coverage has substantial variability across states.^4^

The large-scale distribution of vaccines over a short period across multiple states is a complex logistical problem^5^. While waste is engrained in all production and distribution systems, documenting avoidable vaccine waste that arose during its distribution, including inaccurate demand forecasting, transportation waste, mishandling, improper storage, and expiration, can contribute to preparedness for future, more efficient, and equitable distribution efforts. The Global Alliance for Vaccines and Immunizations (GAVI) recommends at most 25% waste for the first year of the COVID-19 vaccine rollout and 15% by the third year.^6^ Based on these targets, we compare COVID-19 incidence and deaths with counterfactuals representing reduced vaccine wastage and explore three interrelated problems: the variation in COVID-19 vaccine waste across and within states, the implications of vaccine waste towards the prevention of COVID-19 incidence and deaths, and their relations to vaccine hesitancy, urban/rural classification, and political affiliation.

### DATA and METHODS

Doses administered and wastage data by location were obtained from the Centers for Disease Control and Prevention (CDC) through a Freedom of Information Act (case 23-01752). County-level vaccination and hesitancy data were taken from the CDC’s data hub^7^ and the State of Hawaii’s COVID-19 data dashboard,^8^ with additional data obtained from the Census Household Pulse Survey between May 26, 2021, and June 7, 2021 (Supplement eFigure 1).^9^ We also investigate the association between waste, urban/rural classification (Supplement eFigure 2), and political affiliation at the state level (Supplement eFigure 3). We adopt the “red state,” “blue state,” and “purple state” definitions to refer to states whose voters favored the Republican Party, Democratic Party, or a swing state (Supplement eMaterials).

Denoting the number of doses used per capita, *U*_*i*_, and the number of doses wasted per capita, *W*_*i*_, from December 2020 to October 2022. For the *i*^*th*^ county, the percentage of vaccine waste for the *k*^*th*^ state is

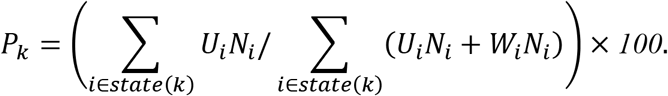

where *i* ∈ *state*(*k*) signifies to add over only the counties in state *k*, and *N*_*i*_ is the population of the *i*^*th*^ county.

For simplicity, we estimate COVID incidence and deaths averted using the Final Size equation^10^ and parameters derived from the literature:

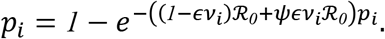

Here, *p*_*i*_ represents the total proportion of the population infected with COVID-19 in the *i*^*th*^ county, *ℛ*_*0*_ = *2*.*3* is the basic reproduction number based on the literature,^11^ *ψ* ≈ *0*.*78* is a reduction in transmission factor caused by vaccination,^12^ *∈* ≈ *0*.*89* is the average efficacy of COVID-19 vaccines,^13^ and *v*_*i*_ is the proportion of the population vaccinated.

We consider interventions where increases in 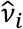, causes counties to reduce waste to 1) 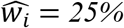 and 2) 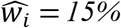. Subsequently, we estimate the proportion infected with COVID-19 under the intervention, 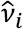, the number of COVID-19 incidences and deaths averted in the *k*^*th*^ state by

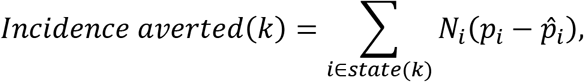

And

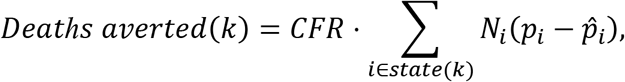

where *CFR* is the case fatality rate of COVID-19, which is 21.8 per 100,000.^14^

## RESULTS

As of October 2022, 761 million vaccine doses were distributed across the U.S. (Supplement eFigure 1). Of these 761 million, 600 million were administered, yielding a national average of 1.8 doses per capita. Despite this average, vaccine uptake varies considerably (Figure 1). Had the U.S. achieved the 25% GAVI target everywhere by increasing uptake, 29,669,318 incidences, and 6,467.9 deaths would have been averted relative to baseline (Table 1). These values increased to 36,130,356 incidences and 7876.4 deaths averted for the 15% GAVI target (Table 1).

**Table 1.**
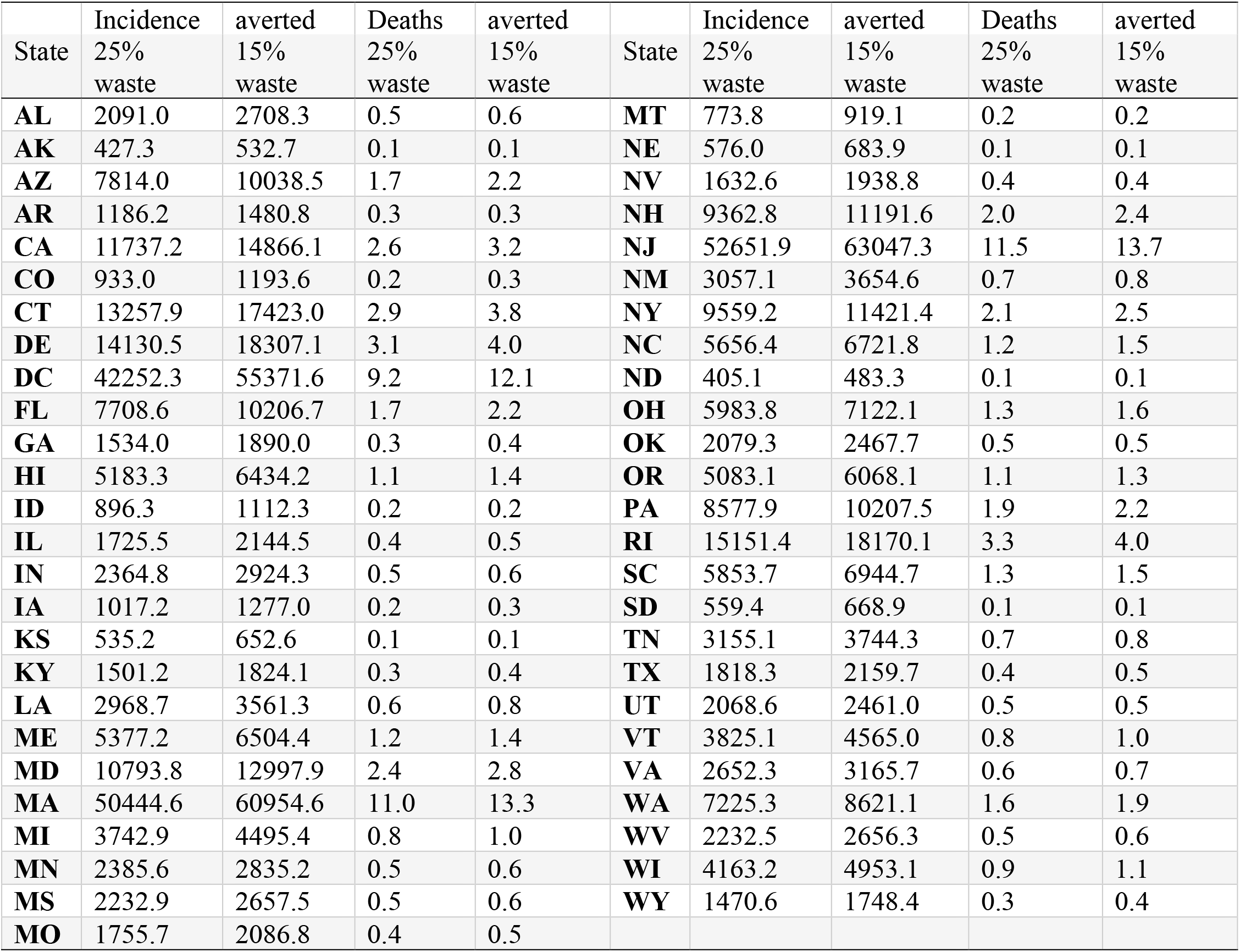
Average incidence and deaths averted for each county exceeding GAVI targets with in a given state.

**Figure 1:**
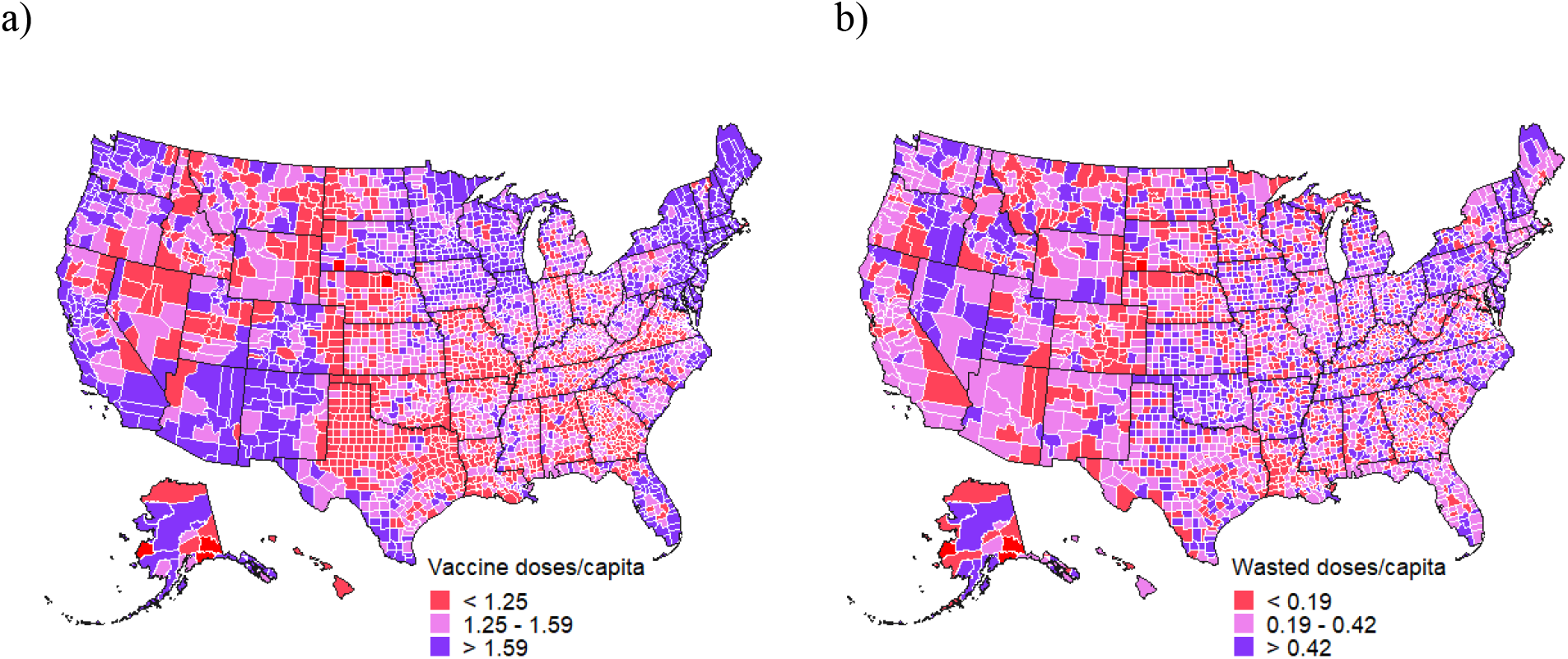
Administered COVID-19 vaccine doses and waste across the U.S. at the county level.

Only Vermont and Michigan exceeded the 25% GAVI target while falling below the average vaccine hesitancy of 10.5% (95% CI: 9.4-11.7) (Figure 2a). Had these states met the 25% GAVI target, 1,072,421 incidences and 23378.79 deaths would have been averted (Table 1, Figure 2b). Nine states exceeded the 15% GAVI target and fell below the average vaccine hesitancy (Figure 2a), which could have averted 8,809,767 incidences, and 192052.9 deaths had the target been met (Table 1, Figure 2b). A total of 21 states exceed the 15% GAVI target and the average vaccine hesitancy (Figure 2a). Most of these states (16 of 21) supported the Republican party in the 2020 elections (Figure 2a). Only 15 states fell below the 15% GAVI target and the average hesitancy rate: 12 blue, one red (South Dakota), and two purple (Maine and Nebraska).

**Figure 2.**
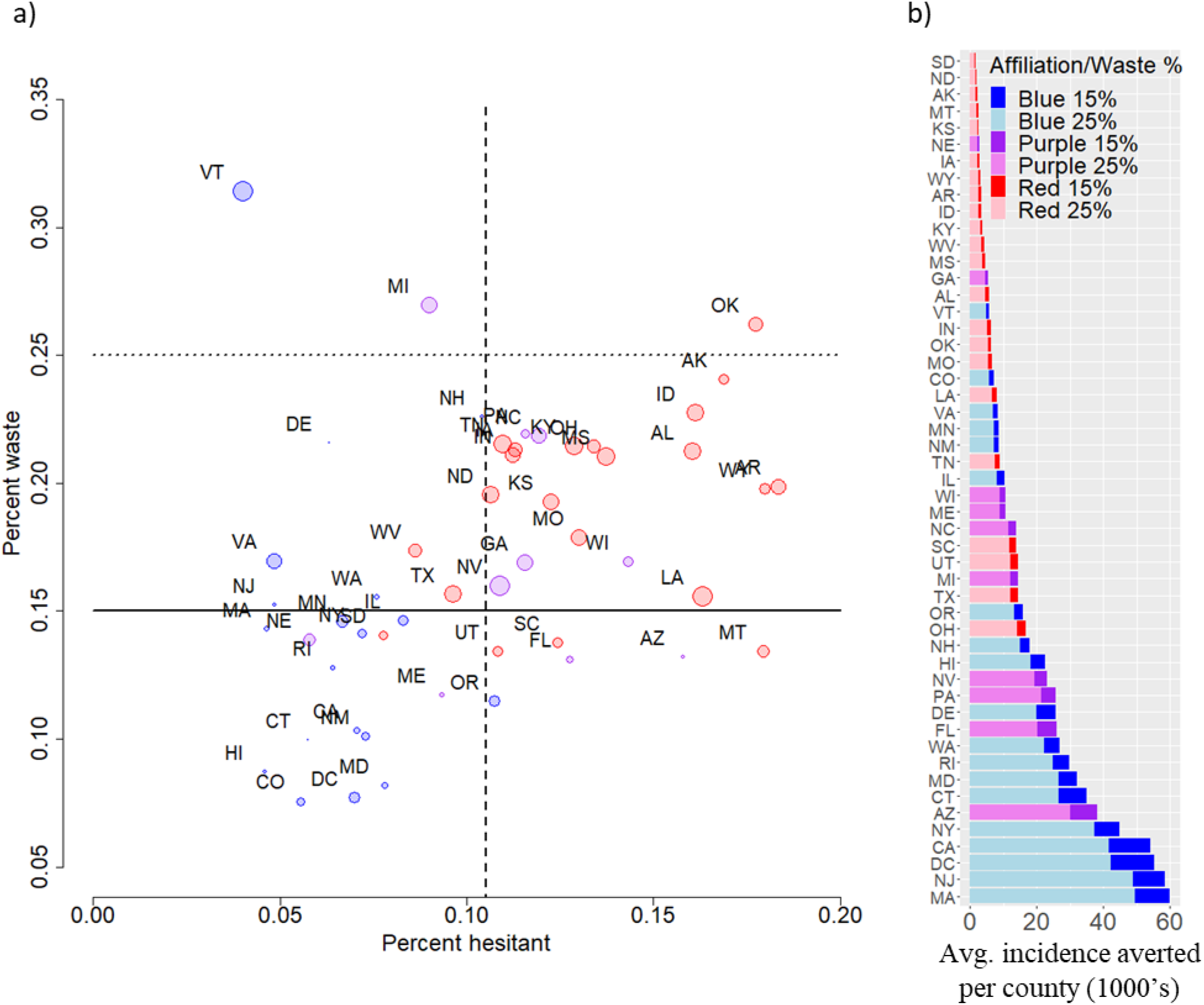
State vaccine waste, hesitancy, and incidence averted. a) The percent of allocated COVID-19 vaccines wasted versus the percentage of the state population reported as vaccine-hesitant. Red, blue, and purple circles represent Republican, Democrat, and swing states from the 2019 presidential elections, where the size of each circle corresponds to the standard deviation in vaccine waste between all counties within the given state (scaled by the standard deviation in vaccine waste across all states). The dotted and solid vertical lines correspond to GAVI waste thresholds of 25% and 15%, respectively, with the solid horizontal line representing the average percentage of people hesitant across all states. b) the average number of COVID-19 incidences averted for each county exceeding the GAVI target within a given state when vaccine waste was reduced to 25% (red 25%, blue 25%, purple 25%) and the additional incidence averted by reducing vaccine waste to 15% (red 15%, blue 15%, and purple 15%), respectively.

At the county level, 901 and 1795 of 3140 exceeded the 25% and 15% GAVI targets, respectively, which amounts to 38.1-64.2 million wasted vaccine doses (Figure 1). This vaccine waste could have averted 0.7-132.6 and 11.7-159.3 incidences per 1000 people per county for the 25% and 15% GAVI targets, respectively. Further separating these numbers by Urban-Rural classification, large central, large fringe, medium, and small metro counties wasted 0.27, 0.354, 0.339, and 0.311 vaccine doses per person, with micropolitan and non-core counties wasting 0.333, and 0.353 vaccine doses per person (Figure 1, Supplement eMethods). Achieving the 25% GAVI target in these metro and non-metro counties would avert between 88.0-90.5 incidences per 1000 people, respectively, with these numbers increasing to 106.3-109.3 incidences per 1000 people for the 15% GAVI target.

## DISCUSSION

While the U.S. at the national level was below the 25% GAVI target in the first 22 months of vaccine administration, the vast discrepancy across counties and states and the predicted incidence and deaths averted highlight the need to improve pandemic preparedness. Simply put, a multi-pronged approach for improvement is required– education and information campaigns for locations with high hesitancy, medical infrastructure improvements in regions with low hesitancy and high waste, and combinations of these two approaches elsewhere.

The divide between Democrats and Republicans strongly correlates with adherence to vaccination recommendations and waste. Towards this regard, a potential avenue for reducing future vaccine waste and hesitancy among Republican counties and states is to examine the particulars of what made South Dakota successful in achieving vaccine waste below 15% and hesitancy below the national average, as replicating such outcomes in other red states could prove vital for future pandemics.

While we inform on domestic vaccine distribution efforts, our work naturally extends to vaccine donation policy. As of 2023, many countries still have less than 10% primary series coverage.^15^ Thus, a moderate decrease in national supply to regions with avoidable waste and subsequent redistribution could substantially impact the global health burden of COVID-19.

This work has limitations. Predictions were based on a simplistic model, with simplifying assumptions placed on epidemiological parameters. Also, vaccine coverage is more complicated than the outright proportion of the population that has completed a vaccine series as the protection by COVID-19 vaccines wanes, although the utilization of multiple boosters conceivably mitigates this effect. These findings thus serve as a benchmark for future, more sophisticated mathematical models based on updated socioeconomic and epidemiological data.

## CONCLUSIONS

Increasing county-level vaccine uptake to achieve the GAVI vaccine waste targets could have substantially reduced COVID-19 incidence and deaths. These findings suggest that future vaccine distribution strategies should consider potential drivers of waste in allocation strategies, as to do so could help increase vaccine uptake, and thereby reduce the negative health outcomes of pandemics.

## Data Availability

All data produced in the present study are available upon reasonable request to the authors.

## ACKNOWLEDGMENTS

We thank the Biomedical Data Science Innovation Lab organization for inviting us to participate in their 2023 summer program and providing the time, space, and support to develop this work. In particular, our appreciation goes to Madhav Marathe and John Van Horn’s insights and encouragement.

## Supplemental materials

These supplemental materials provide further details on the mathematical methods used to estimate the proportions of vaccines wasted, along with illustrations and sources of data for vaccine hesitancy, Urban-Rural classification, and political affiliation used in the analysis.

## S1. Mathematical methods

For each county, we calculate the proportion of vaccine waste as

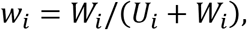

and compute the variance in the proportion of vaccine waste over all counties within the *k*^*th*^ state as

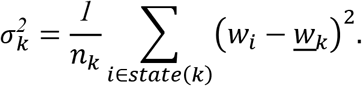

Here *n*_*k*_ represents the number of counties and *w*_*k*_ is the average proportion of vaccine waste in the *k*^*th*^ state.

### S2. Vaccine hesitancy

To determine the percentage of each county that is COVID-19 vaccine-hesitant, we obtained survey data from the Census Household Pulse Survey between May 26, 2021, and June 7, 2021 (Figure S1).^1,2^

**eFigure 1.**
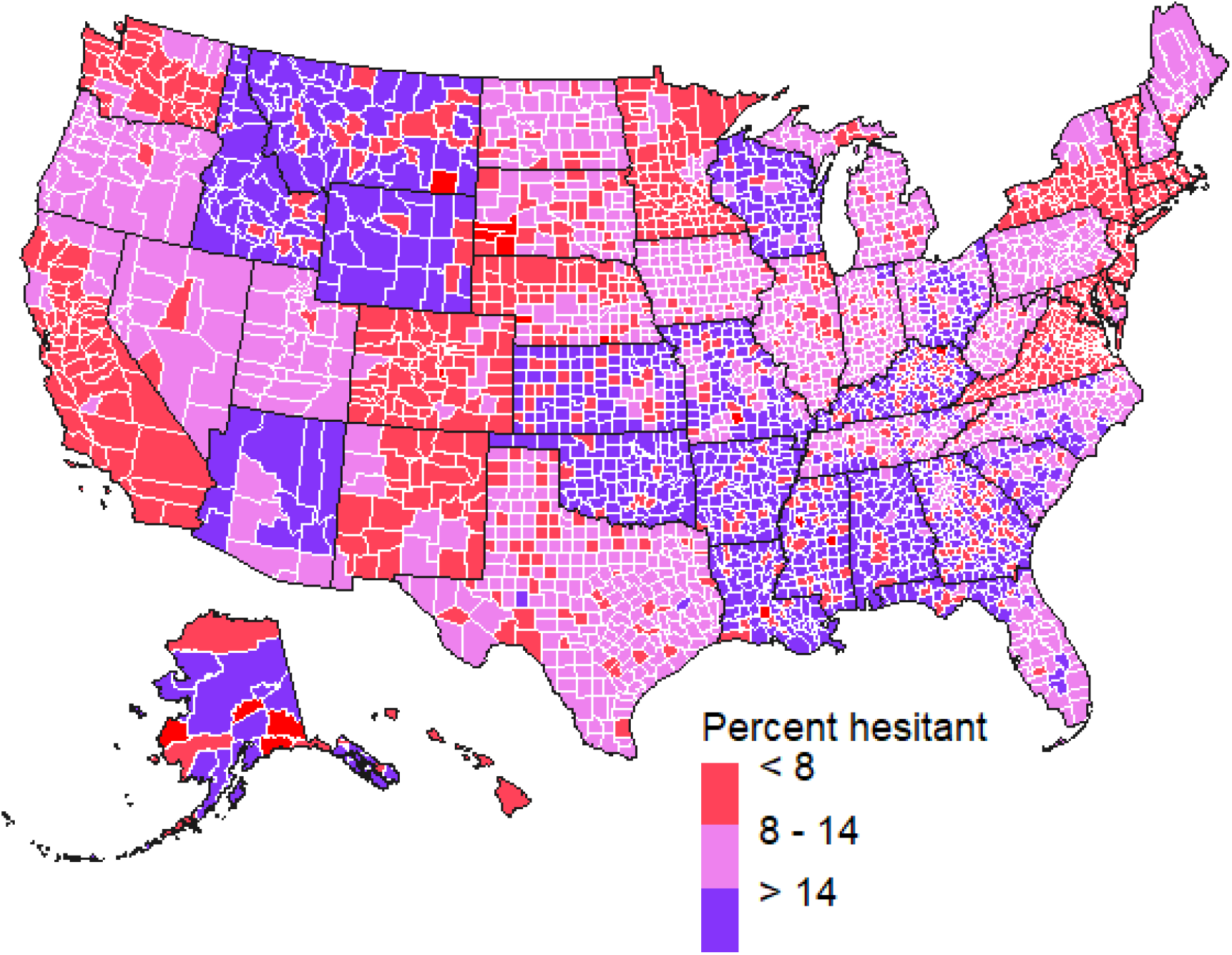
Percentage of each county that is COVID-19 vaccine-hesitant.

### S3. Urban-Rural classification

We consider the National Center for Health Statistics classification of urban-rural counties^3^. Roughly, counties are grouped into metropolitans as either 1) large central metro, 2) large fringe metro, 3) medium metro, 4) small metro, or into a nonmetropolitan as 5) micropolitan or 6) non-core (Figure S2).

**eFigure 2.**
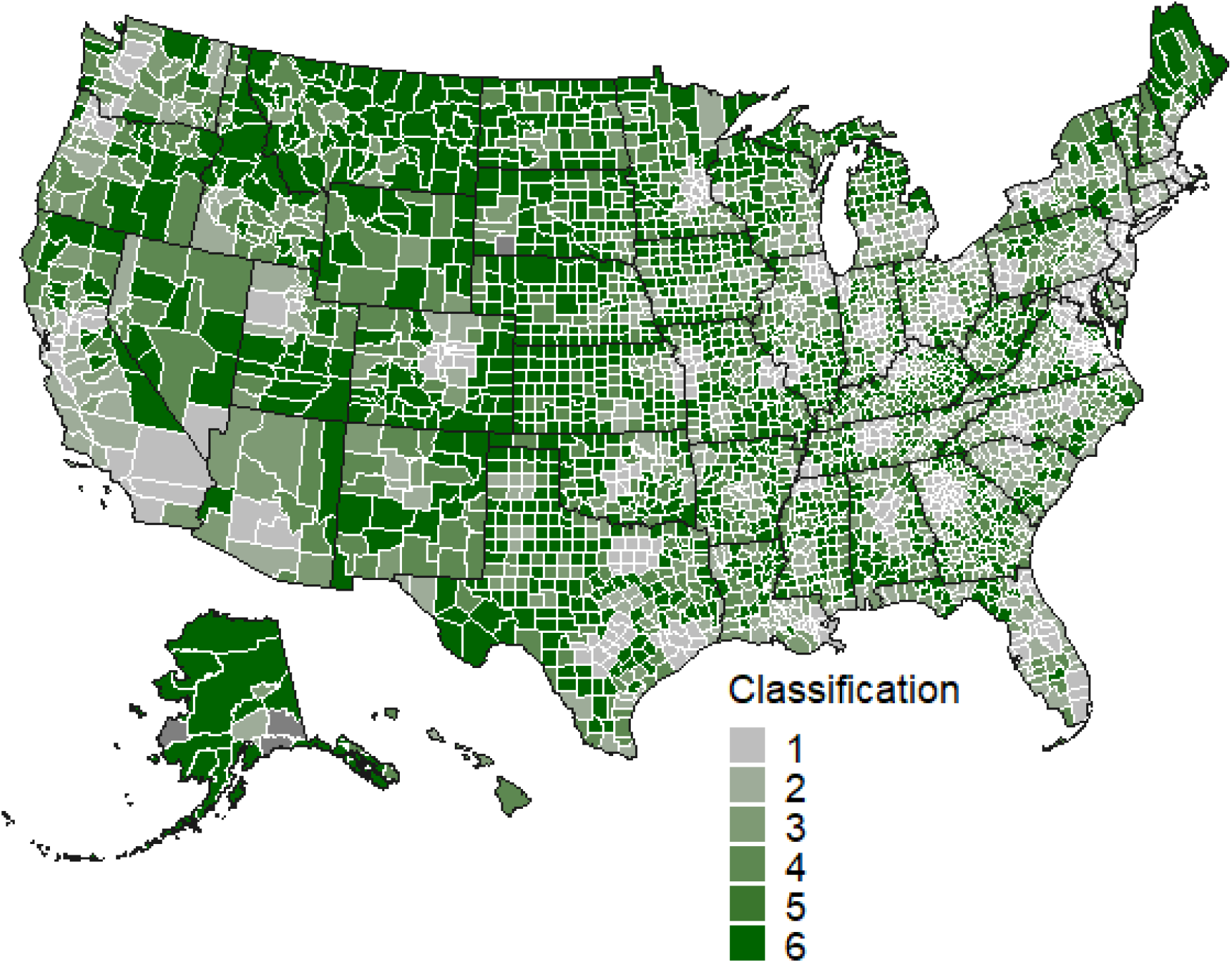
Urban-rural classification of counties. The classification of 1 (grey) to 6 (dark green) corresponds to counties classified as large central metro, large fringe metro, medium metro, small metro, micropolitan, and non-core, respectively.

### S4. Political affiliation

We adopt the “red state” and “blue state” definitions to refer to states whose voters favored the Republican Party and the Democratic Party with a margin higher than 5%, respectively, in the 2020 presidential elections, with other states referred to as “purple states”.^4^

**eFigure 3.**
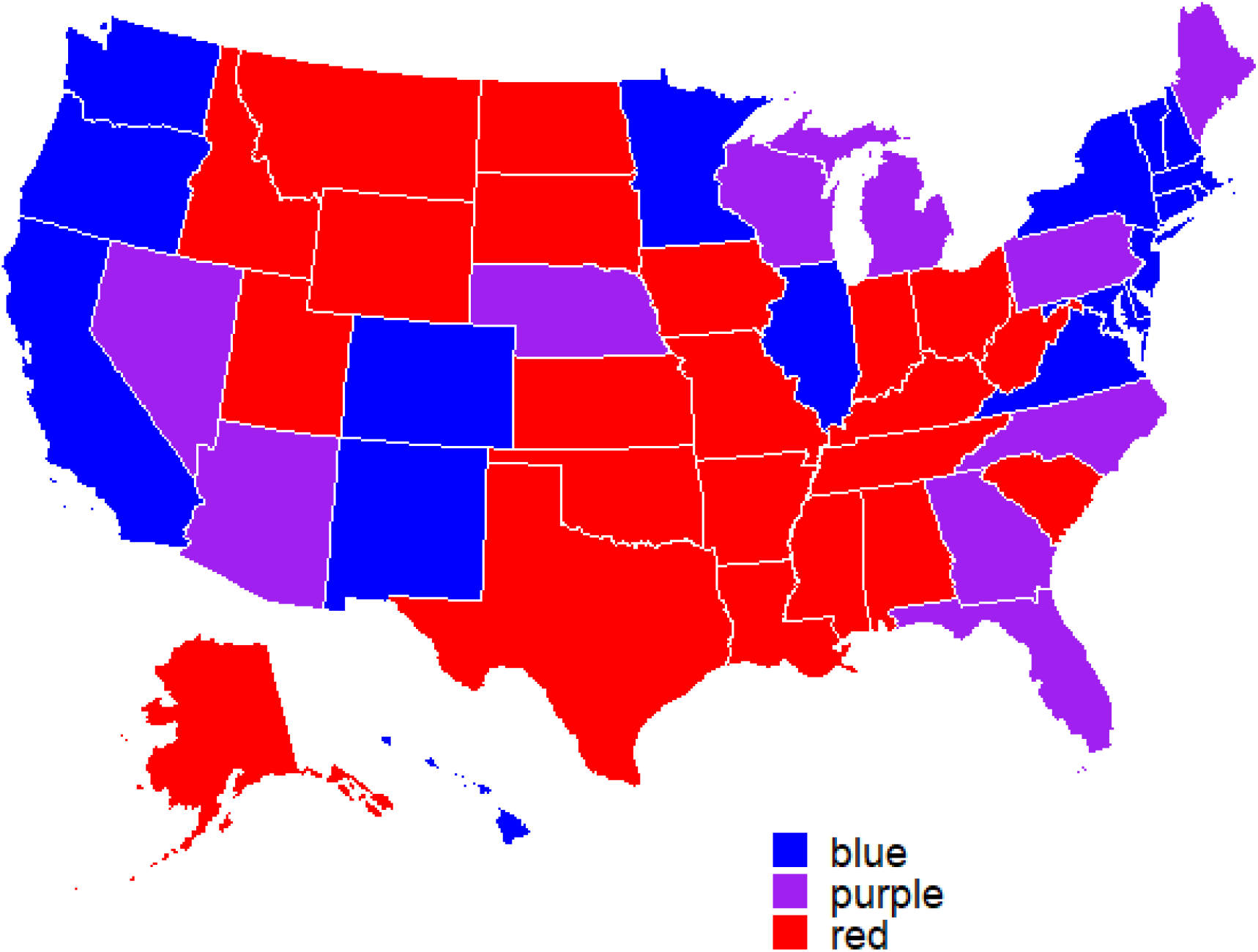
Urban-rural classification of counties.

